# Trends in youth use of non-cigarette tobacco products in England, Canada, and the US and the impact of England’s menthol cigarette ban on use

**DOI:** 10.1101/2025.11.12.25340079

**Authors:** Yamma Khalid Aria, Sarah E. Jackson, David Hammond, Jessica L. Reid, Eve Taylor, Madeleine Ebdon, Harry Tattan-Birch, Masuma Mishu, Jamie Brown, Lion Shahab, Sharon Cox, Katherine East

## Abstract

**Introduction:** In England, non-cigarette tobacco products (e.g., cigarillos, cigars, waterpipes, bidis, smokeless tobacco, heated tobacco) are subject to fewer regulatory restrictions than cigarettes/rolling tobacco. This study assessed trends in youth use of these products in England, Canada, and the US, focussing on evaluating the impact of England’s May 2020 ban on menthol in cigarettes but not non-cigarette tobacco.

**Methods:** Data were from the ITC Youth Tobacco and Vaping Survey (16-19-year-olds; N=129,575) across ten waves (2017–2024). Segmented regressions assessed changes in trends of non-cigarette tobacco products before vs. after May 2020, adjusting for race/ethnicity/sex/age.

**Results:** In England, non-cigarette tobacco use increased similarly before (aOR=1.16,1.02–1.31) and after (1.13,1.07–1.20) the ban (change in trend: 0.98,0.91–1.06), from 7.4% in 2017 and reaching in 11.6% in 2024. In contrast, non-cigarette tobacco use decreased similarly before and after the ban in both the US (pre-ban: 0.82, 0.73–0.93; post-ban: 0.84,0.80–0.89; change in trend: 1.02,0.94–1.11; from 10.2% reaching 7.1%), and Canada (pre-ban: 0.84,0.76–0.93; post-ban: 0.90,0.86–0.94; from 9.2% reaching 7.9%), although the decline in Canada after the ban was slightly slower (1.07,1.001–1.15). Cigarillo, cigar, and smokeless tobacco use specifically increased to a greater extent in England than Canada and the US.

**Conclusions:** Between 2017 and 2024, youth non-cigarette tobacco product use increased in England and surpassed declining use in Canada and the US. There was little evidence that England’s menthol cigarette ban accelerated increases, although increases highlight the need for comprehensive regulations encompassing all tobacco products in England.

**IMPLICATIONS:** Youth use of non-cigarette tobacco products has been increasing in England but declining in Canada and the US. England’s menthol cigarette ban did not appear to alter trends in use of non-cigarette tobacco, but findings highlight a need for more comprehensive regulation encompassing all tobacco products to mitigate youth uptake.

## INTRODUCTION

Tobacco smoking remains a leading cause of cancer and premature death.^1^ While cigarette smoking is the most prevalent form of tobacco use in England, an estimated 3.7% of adults (∼1.7 million people) currently use non-cigarette tobacco products.^2^ This includes combustible options such as cigars, cigarillos, waterpipes (shisha), bidis, and pipes, as well as non-combustible options like snuff, chewing tobacco, heated tobacco products, and snus (not legally sold in the United Kingdom (UK)). All tobacco use carries risks; however, combustible products generally pose greater risks.^3^

According to the Smoking Toolkit Study, in 2024/2025, after cigarettes, cigars were the most commonly used tobacco product among adults in England, followed by waterpipe/shisha, cigarillos, pipes, and bidis.^2^ Non-combustible tobacco products were less common, with snus, heated tobacco, and other smokeless tobacco each used by fewer than 1% of adults.^2^ Exclusive use of non-cigarette tobacco products (without cigarettes) rose from 1.3% in 2020 to 2.0% in 2022,^2^ although this since declined to 1.2% in early 2025.^2^ This aligns with Her Majesty Revenue and Customs (HMRC) figures showing UK cigar sales increasing from £73 million in 2019 to £165 million in 2024.^4^ Less is known about youth use.

In the UK, some policies differ between cigarettes/rolling tobacco and non-cigarette tobacco products. For example, while menthol and other flavours in cigarettes were prohibited in May 2020, they remain available in non-cigarette tobacco products. Further, standardised packaging and a minimum pack size for cigarettes (20 cigarettes) and rolling tobacco (30g) was implemented in May 2017; however, this does not apply to non-cigarette tobacco products, which can be purchased in colourful, branded packaging, and smaller packs (e.g., cigars are often sold singly, cigarillos are often sold in packs of 10). Flavours, colourful packaging, and lower costs increase the appeal and use of tobacco products, particularly among youth.^5–8^ Therefore, excluding non-cigarette tobacco products from these policies may risk substitution of cigarettes with non-cigarette tobacco. This risk is exacerbated by the tobacco industry’s response to UK legislation, such as introducing flavour/capsule ‘cigarette-like-cigarillos’ and promoting flavoured heated tobacco sticks around the time menthol-flavoured cigarettes were banned.^9^

Unlike the UK, Canada prohibits menthol/flavours in blunt wraps and most cigars/cigarillos as well as cigarettes. Canada federally prohibited most flavours in January 2016, followed by the menthol in October 2017,^10^ although several provinces introduced regulations earlier. Also, unlike the UK, Canada implemented standardised packaging of all tobacco products in February 2020^11^ and requires minimum pack sizes of 20 for cigarettes, little cigars, and blunt wraps, but not other tobacco products (e.g., regular cigars). Unlike both the UK and Canada, the United States (US) does not currently have a federal ban on flavours in tobacco products (cigarette or non-cigarette) or on standardised packaging/pack sizes, but did raise the legal age of sale for all tobacco products from 18 to 21 in December 2019.^12^ Several US jurisdictions (e.g., California, Massachusetts, New Jersey, New York) also implemented banned menthol cigarettes and flavoured non-cigarette tobacco products.

The variation in policies across England, Canada, and the US, specifically England’s ban on menthol and other flavours in cigarettes in May 2020, creates conditions for a natural experiment. This study therefore uses a quasi-experimental design to address the following research questions:

1. Has the prevalence of youth use of non-cigarette tobacco products (any, cigarillos, cigars, waterpipes, bidis, smokeless tobacco, and heated tobacco) changed between 2017 and 2024 in each of England, Canada and the US?
2. Have trends in the prevalence of youth use of non-cigarette tobacco products over this period differed between England, Canada, and the US?
3. Has England’s ban on menthol in cigarettes (but not other tobacco products) impacted prevalence of non-cigarette tobacco use?

Hypotheses are:

1. In England, prevalence of youth use of the following non-cigarette tobacco products will have increased after May 2020: any, cigarillos, cigars, heated tobacco.
2. Youth use of these products (any, cigarillos, cigars, heated tobacco) will have increased more in England compared with Canada and the US.
3. After England’s menthol cigarette ban, the proportion of youth using menthol (vs. non-menthol) non-cigarette products (any, cigarillos, cigars, heated tobacco) will be relatively higher in England than in Canada or the US.

## METHODS

Analyses were pre-registered and the code is available online (https://osf.io/r2y54).^13^

### Data source

Data were from the International Tobacco Control Project (ITC) Youth Tobacco and Vaping Survey, a repeated cross-sectional online survey of youth aged 16-19 in England, Canada, and the US, conducted annually or twice annually. Ten survey waves were conducted from 2017 through 2024. Study methods are described online.^14^ A total of N=129,575 respondents were retained in the analytic sample. This study was reviewed and received ethics clearance through a University of Waterloo Research Ethics Committee (ORE#21847/31017).

### Measures

Measure details are available on the Open Science Framework (https://osf.io/r2y54).^13^ Primary outcomes were past 30-day use (yes vs. other) of (a) any non-cigarette tobacco product (b-g and, for sensitivity analysis, b-f), (b) little cigars/cigarillos, (c) cigars, (d) bidis, (e) smokeless tobacco, (f) waterpipe/shisha, and, from 2018 onwards, (g) heated tobacco. For use of each product, respondents could select ‘yes’, ‘no’, or ‘don’t know’; responses of ‘no’ and ‘don’t know’ were combined into the ‘other’ category for analysis.

Secondary outcomes were past 30-day use (yes vs. other, assessed from 2021 onwards) of menthol-flavoured (a) any non-cigarette tobacco product (b-g and, for sensitivity analysis, b-f), (b) little cigars/cigarillos, (c) cigars, (d) bidis, (e) smokeless tobacco, (f) waterpipe/shisha, and (g) heated tobacco. As above, for each product, respondents could select ‘yes’, ‘no’, or ‘don’t know’; the latter two were combined into ‘other’.

Key predictors were country (England, Canada, US) and survey wave (Jul/Aug 2017, Aug/Sept 2018, Aug/Sept 2019, Feb/Mar 2020, Aug 2020, Feb/Mar 2021, Aug/Sept 2021, Aug/Sept 2022, Aug/Sept 2023, Aug/Sept 2024). Survey wave was first treated as categorical to assess wave-by-wave changes. Then, survey wave was treated as continuous to assess overall trends, with a dummy variable created to indicate the pre-May 2020 period and post-May 2020 period (i.e., before vs. after England’s menthol cigarette ban).

Sociodemographic covariates were race/ethnicity (white, black, other, multiple, don’t know/refused), sex-at-birth (male, female), and age (16-17, 18-19 years).

### Sample weighting

Cross-sectional survey weights were used in all analyses to enhance representativeness of the data to the populations from which the samples were derived.

Briefly, cross-sectional poststratification sample weights were constructed for each country baed on population figures for sex-by-age-by-region (sex-by-age-by-region-by race in the US), calibrated to wave 1 student status and school grades and, in Canada and the US only, past 30-day smoking trend, and rescaled to each country’s sample size. Further details are available online.^14^

### Statistical analyses

For Research Questions 1 and 2, we reported the weighted proportions (and 95% Confidence Intervals [CIs]) per country and survey wave for each primary outcome. We then used logistic regression models with each primary outcome and country, survey wave (categorical), and a country*survey wave interaction term as predictors, adjusting for sociodemographic covariates. Average adjusted probabilities were estimated from these models and contrasted within and between countries.

For Research Question 3, we used segmented regression analyses (using binary logistic regressions) to assess whether England’s menthol cigarette ban impacted trends in the prevalence of non-cigarette tobacco product use. We modelled trends for each primary outcome before the interruption (underlying secular trend; coded 0 … n, where n was the total number of waves scaled to account for timing between waves, e.g., 0.0, 0.5, 1.0, 1.25…

3.5 representing Jul/Aug 2017, Aug/Sep 2018, Aug/Sep 2019, Feb/Mar 2020… Aug/Sep 2024) and the change in the trend (slope) post-ban relative to pre-ban (coded 0 before May 2020 and 0.25 … m from May 2020 onwards, again scaled for timing between waves). Models were adjusted for country, sociodemographic covariates, and country*change and country*trend interaction term. Adjusted estimates were then generated and contrasted within and between countries.

To assess any past 30-day use of menthol (vs. exclusively non-menthol) non-cigarette tobacco products (i.e., secondary outcomes) among youth reporting past 30-day use of the tobacco product being assessed, we first reported proportions in each country, aggregating data from August 2021 onwards (when this question was first included in the survey, after England’s May 2020 menthol cigarette ban). Second, logistic regression models were used with each secondary outcome and predictors listed above. Average adjusted probabilities were then estimated and contrasted within and between countries.

Analyses used STATA v18.

### Sensitivity analysis

Analyses were repeated excluding heated tobacco from the primary and secondary outcomes because heated tobacco was not assessed in 2017 and IQOS (a leading brand of heated tobacco) was removed from the US market in November 2021. Prevalence of past 30-day menthol (vs. non-menthol) cigarette use at each wave is reported in the supplementary materials for comparison.

## RESULTS

### Sample characteristics

Table 1 shows the sample characteristics. Following weighting, most participants identified as white and there was a relatively equal split of males/females and age groups.

**Table 1.** Demographic characteristics by country.

| Characteristics |  | Canada<br>(n=42,499) | England<br>(n=41,327) | United States<br>(n=45,749) |
| --- | --- | --- | --- | --- |
| Sex at birth | Male | 51.2% (16,083) | 51.4% (15,166) | 51.1% (14,650) |
|  | Female | 48.8% (26,416) | 48.6% (26,161) | 48.9% (31,099) |
| Age group<br>(years) | 16–17 | 47.9% (18,969) | 49.3% (17,134) | 49.6% (22,006) |
|  | 18–19 | 52.1% (23,530) | 50.7% (24,193) | 50.40% (23,743) |
| Race/Ethnicity<sup>1</sup> | White | 54.4% (23,535) | 65.7% (26,677) | 64.9% (22,494) |
|  | Black | 6.2% (2,701) | 5.9% (2,614) | 7.8% (5,699) |
|  | Other | 28.2% (11,363) | 11.5% (4,859) | 9.9% (7,214) |
|  | Multiple | 8.5% (3,660) | 6.2% (2,701) | 7.9% (5,840) |
|  | Don't know/refused | 2.7% (1,240) | 10.7% (4,476) | 9.5% (4,502) |

*Note: Data are weighted % (unweighted n).*
<sup>1</sup> *Derived variable based on country-specific measures.*

### Prevalence, trends, and country differences in youth use of non-cigarette tobacco products (Research Questions 1 and 2)

#### Within-country time trends

In England, use of any non-cigarette tobacco product increased from 7.4% in 2017 to 11.6% in 2024 (aOR=2.22,95% CI=1.78-2.79; Figure 1; Table 2). Use rose specifically from 2017 to Feb/Mar 2020 (aOR=2.13,1.70-2.67) before subsequently dropping through to Feb/Mar 2021 (vs. Feb/Mar 2020; aOR=0.69,0.58-0.82), then increasing again in 2024 (vs. Feb/Mar 2021; 8.4% to 11.6%; aOR=1.52,1.29-1.80) (data not shown in tables).

**Figure 1.**
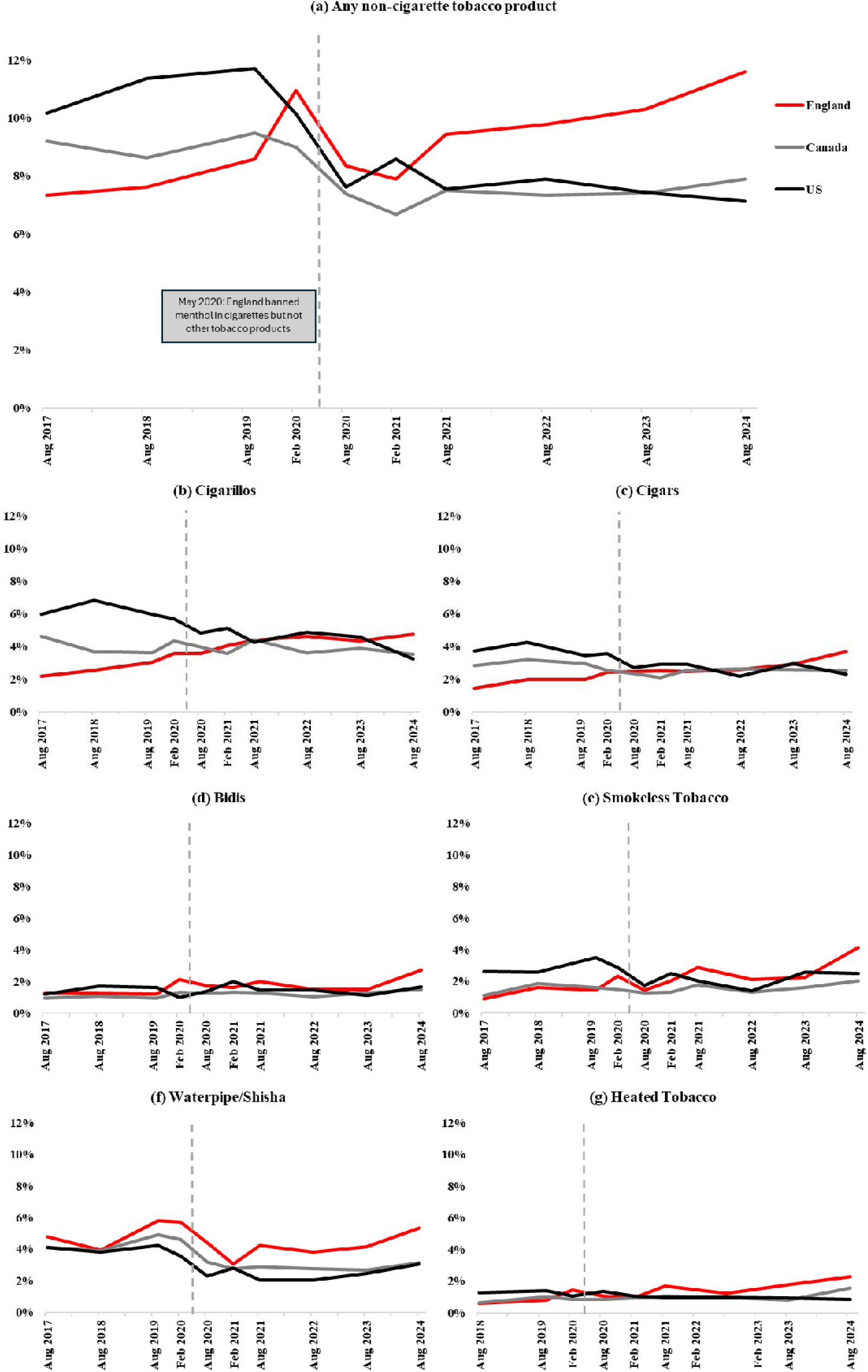
Prevalence of past 30-day use of non-cigarette tobacco products among youth aged 16-19 in Canada, England, and the US between 2017 and 2024. **Alt Text. Graphs showing general increases in use of non-cigarette tobacco products in England alongside**

**Table 2.** Changes over time in the proportion of youth (16-19 years) who reported past 30-day use of any non-cigarette tobacco product within England, Canada, and the US.

|  | England (n=41,327) |  |  | Canada (n=42,499) |  |  | United States (n=45,749) |  |  |
| --- | --- | --- | --- | --- | --- | --- | --- | --- | --- |
|  | %(n) | aOR (95% CI) | P | %(n) | aOR (95% CI) | P | %(n) | aOR (95% CI) | P |
| **Any use of non-cigarette tobacco products** |  |  |  |  |  |  |  |  |  |
| 2017 (Jul-Aug) | 7.4 (306) | **REF** | — | 9.2 (334) | **REF** | — | 10.2 (412) | **REF** | — |
| 2018 (Aug-Sep) | 7.6 (286) | **1.43 (1.12 - 1.82)** | **.004** | 8.6 (436) | 0.90 (0.76 - 1.06) | .214 | 11.4 (443) | 1.52 (1.22 - 1.91) | **<.001** |
| 2019 (Aug-Sep) | 8.6 (328) | **1.62 (1.28 - 2.05)** | **<.001** | 9.5 (450) | 1.00 (0.84 - 1.18) | .962 | 11.7 (578) | 1.57 (1.25 - 1.98) | **<.001** |
| 2020 (Feb-Mar) | 11.0 (511) | **2.13 (1.70 - 2.67)** | **<.001** | 9.0 (474) | 0.94 (0.79 - 1.11) | .472 | 10.2 (633) | **1.33 (1.07 - 1.67)** | **.012** |
| 2020 (Aug) | 8.4 (395) | **1.57 (1.24 - 1.98)** | **<.001** | 7.4 (295) | **0.77 (0.64 - 0.91)** | **.003** | 7.6 (496) | 0.97 (0.77 - 1.22) | .796 |
| 2021 (Feb-Mar) | 7.9 (327) | **1.46 (1.15 - 1.86)** | **.002** | 6.7 (296) | **0.69 (0.57 - 0.82)** | **<.001** | 8.6 (507) | 1.09 (0.86 - 1.38) | .477 |
| 2021 (Aug-Sep) | 9.4 (440) | **1.78 (1.42 - 2.25)** | **<.001** | 7.5 (339) | **0.78 (0.65 - 0.93)** | **.005** | 7.6 (455) | 0.96 (0.75 - 1.23) | .761 |
| 2022 (Aug-Sep) | 9.8 (480) | **1.88 (1.50 - 2.36)** | **<.001** | 7.3 (360) | **0.75 (0.63 - 0.90)** | **.002** | 7.9 (403) | 1.02 (0.79 - 1.31) | .891 |
| 2023 (Aug-Sep) | 10.3 (478) | **1.96 (1.56 - 2.47)** | **<.001** | 7.4 (345) | **0.77 (0.64 - 0.91)** | **.003** | 7.4 (391) | 0.95 (0.74 - 1.22) | .672 |
| 2024 (Aug-Sep) | 11.6 (527) | **2.23 (1.78 - 2.79)** | **<.001** | 7.9 (358) | **0.82 (0.68 - 0.98)** | **.028** | 7.1 (454) | 0.90 (0.70 - 1.15) | .407 |
| 2023 (Aug-Sep) | 4.2 (188) | 0.97 (0.68 - 1.37) | .855 | 2.7 (119) | **0.64 (0.49 - 0.84)** | **.001** | 2.5 (121) | **0.66 (0.44 - 1.000)** | **.050** |
| 2024 (Aug-Sep) | 5.3 (236) | 1.25 (0.89 - 1.76) | .189 | 3.1 (138) | **0.75 (0.57 - 0.98)** | **.033** | 3.1 (170) | 0.83 (0.56 - 1.22) | .338 |

*Note: All data except for n are weighted. aOR, 95% CIs, and p values were derived using Stata's post-estimation margins command after fitting separate logistic regression models (one for each outcome), including a survey wave by country interaction term and adjusting for age group, sex, and race/ethnicity. 95% CIs are presented to two decimal places except where they are close to 1, in which case three decimal places are reported. Values in bold indicate p-values <.05.*

In Canada, there was modest evidence of an overall decrease between 2017 and 2024 (9.2% to 7.9%, aOR=0.82,0.68-0.98), although the decrease was most pronounced between 2017 and Feb/Mar 2021 (9.2% to 6.7%, aOR=0.69, 0.57-0.82). In the US, prevalence decreased between 2017 and 2024 (10.2% to 7.1%) although when adjusting for covariates there was little evidence of a difference (aOR=0.90,0.70-1.15) (Figure 1; Table 2).

For individual products, in England, between 2017 and 2024, there were consistent modest increases in past-30-day use of most products (Figure 1; Tables 3 and 4). Use of cigarillos rose from 2.2% to 4.7%, cigars from 1.4% to 3.7%, bidis from 1.2% to 2.7%, smokeless tobacco from 0.9% to 4.1%, and heated tobacco from 0.6% in 2018 to 2.3% (all p<.001; Tables 3 and 4). Notably, modest increases from August 2020 to 2024 – the period after the menthol ban – were evident for cigarillos (3.6% to 4.7%; aOR=1.32,1.03–1.69), cigars (2.4% to 3.7%; aOR=1.53,1.15–2.04), bidis (2.1% to 2.7%; aOR=1.53,1.09–2.14), smokeless tobacco (1.4% to 4.1%; aOR=3.01,2.15–4.20), and heated tobacco (1.1% to 2.3%; aOR=2.18,1.49–3.21) (data not shown in tables). While overall waterpipe use in England remained relatively stable from 2017 to 2024 (4.8% to 5.3%; Table 4), there was an increase from Feb/Mar 2021 to 2024 (3.1% to 5.3%; aOR=1.76,1.36–2.28; data not shown in tables). In contrast, in Canada, cigarillo use showed modest evidence of decline between 2017 and 2024 (4.6% to 3.5%), waterpipe use declined somewhat (4.1% to 3.1%), cigars remained stable (2.8% to 2.5%), while smokeless (1.1% to 2.0%) and heated (0.6% in 2018 to 1.6%) tobacco use increased (Tables 3 and 4). The US showed declines or stability between 2017 and 2024 in use of cigarillos (6.1% to 3.2%), cigars (3.7% to 2.3%), waterpipe (4.1% to 3.1%), and heated tobacco (1.3% to 0.8%), while use of bidis increased slightly despite fluctuations over the study period (1.2% to 1.7%) (Tables 3 and 4).

**Table 3.** Changes over time in the proportion of youth (16-19 years) who reported past 30-day use of cigarillos, cigars, and bidis within England, Canada, and the US.

|  | England (n=41,327) |  |  | Canada (n=42,499) |  |  | United States (n=45,749) |  |  |
| --- | --- | --- | --- | --- | --- | --- | --- | --- | --- |
|  | %(n) | aOR (95% CI) | P | %(n) | aOR (95% CI) | P | %(n) | aOR (95% CI) | P |
| **Cigarillos** |  |  |  |  |  |  |  |  |  |
| 2017 (Jul-Aug) | 2.2 (81) | REF | — | 4.6 (153) | REF | — | 6.0 (232) | REF | — |
| 2018 (Aug-Sep) | 2.5 (87) | 1.26 (0.83 - 1.92) | .270 | 3.7 (191) | **0.78 (0.62 - 0.99)** | **.045** | 6.8 (264) | 1.22 (0.87 - 1.70) | .250 |
| 2019 (Aug-Sep) | 3.0 (111) | **1.52 (1.02 - 2.28)** | **.042** | 3.6 (172) | **0.73 (0.57 - 0.94)** | **.015** | 6.0 (320) | 1.05 (0.75 - 1.47) | .773 |
| 2020 (Feb-Mar) | 3.5 (157) | **1.77 (1.20 - 2.62)** | **.004** | 4.3 (211) | 0.89 (0.70 - 1.14) | .353 | 5.7 (377) | 1.00 (0.71 - 1.39) | .977 |
| 2020 (Aug) | 3.6 (162) | **1.78 (1.21 - 2.64)** | **.004** | 4.0 (148) | 0.82 (0.64 - 1.06) | .127 | 4.8 (294) | 0.84 (0.60 - 1.18) | .325 |
| 2021 (Feb-Mar) | 4.0 (149) | **2.03 (1.37 - 3.02)** | **<.001** | 3.6 (149) | **0.73 (0.56 - 0.94)** | **.016** | 5.1 (308) | 0.89 (0.63 - 1.26) | .502 |
| 2021 (Aug-Sep) | 4.4 (181) | **2.22 (1.49 - 3.30)** | **<.001** | 4.4 (183) | 0.92 (0.72 - 1.17) | .495 | 4.3 (261) | 0.74 (0.52 - 1.06) | .105 |
| 2022 (Aug-Sep) | 4.6 (225) | **2.36 (1.62 - 3.45)** | **<.001** | 3.6 (177) | **0.73 (0.57 - 0.94)** | **.014** | 4.9 (239) | 0.85 (0.59 - 1.23) | .390 |
| 2023 (Aug-Sep) | 4.4 (217) | **2.20 (1.49 - 3.24)** | **<.001** | 3.9 (180) | 0.81 (0.63 - 1.03) | .086 | 4.6 (240) | 0.80 (0.56 - 1.15) | .226 |
| 2024 (Aug-Sep) | 4.7 (196) | **2.36 (1.61 - 3.46)** | **<.001** | 3.5 (166) | **0.72 (0.56 - 0.93)** | **.012** | 3.2 (253) | **0.55 (0.38 - 0.79)** | **.001** |
| **Cigars** |  |  |  |  |  |  |  |  |  |
| 2017 (Jul-Aug) | 1.4 (54) | REF | — | 2.8 (95) | REF | — | 3.7 (146) | REF | — |
| 2018 (Aug-Sep) | 2.0 (67) | **2.71 (1.73 - 4.26)** | **<.001** | 3.2 (170) | 1.07 (0.81 - 1.41) | .631 | 4.3 (147) | **2.21 (1.56 - 3.15)** | **<.001** |
| 2019 (Aug-Sep) | 2.0 (69) | **2.70 (1.74 - 4.20)** | **<.001** | 2.9 (143) | 1.00 (0.74 - 1.34) | .984 | 3.4 (164) | **1.77 (1.24 - 2.54)** | **.002** |
| 2020 (Feb-Mar) | 2.4 (109) | **3.34 (2.20 - 5.08)** | **<.001** | 2.5 (122) | 0.86 (0.63 - 1.17) | .347 | 3.5 (221) | **1.83 (1.29 - 2.60)** | **.001** |
| 2020 (Aug) | 2.4 (106) | **3.39 (2.23 - 5.16)** | **<.001** | 2.3 (81) | 0.79 (0.57 - 1.10) | .167 | 2.7 (171) | 1.40 (0.97 - 2.00) | .070 |
| 2021 (Feb-Mar) | 2.5 (88) | **3.49 (2.26 - 5.41)** | **<.001** | 2.1 (91) | **0.71 (0.51 - 0.99)** | **.041** | 2.9 (160) | **1.49 (1.02 - 2.18)** | **.038** |
| 2021 (Aug-Sep) | 2.5 (106) | **3.46 (2.24 - 5.34)** | **<.001** | 2.6 (106) | 0.88 (0.64 - 1.20) | .409 | 2.9 (159) | **1.50 (1.01 - 2.22)** | **.043** |
| 2022 (Aug-Sep) | 2.6 (115) | **3.61 (2.37 - 5.51)** | **<.001** | 2.6 (119) | 0.89 (0.66 - 1.22) | .478 | 2.2 (133) | 1.11 (0.75 - 1.64) | .603 |
| 2023 (Aug-Sep) | 2.9 (120) | **4.12 (2.71 - 6.25)** | **<.001** | 2.6 (111) | 0.89 (0.66 - 1.22) | .477 | 2.9 (145) | **1.52 (1.03 - 2.24)** | **.037** |
| 2024 (Aug-Sep) | 3.7 (154) | **5.20 (3.47 - 7.79)** | **<.001** | 2.5 (115) | 0.87 (0.63 - 1.20) | .388 | 2.3 (176) | 1.18 (0.81 - 1.73) | .397 |
| **Bidis** |  |  |  |  |  |  |  |  |  |
| 2017 (Jul-Aug) | 1.2 (48) | REF | — | 1.0 (29) | REF | — | 1.2 (43) | REF | — |
| 2018 (Aug-Sep) | 1.2 (42) | 1.73 (0.98 - 3.07) | .060 | 1.1 (60) | 1.06 (0.65 - 1.72) | .818 | 1.7 (63) | **2.45 (1.39 - 4.32)** | **.002** |
| 2019 (Aug-Sep) | 1.2 (52) | 1.69 (0.97 - 2.93) | .064 | 0.9 (53) | 0.95 (0.57 - 1.57) | .838 | 1.6 (75) | **2.31 (1.29 - 4.13)** | **.005** |
| 2020 (Feb-Mar) | 2.1 (98) | **3.03 (1.80 - 5.11)** | **<.001** | 1.3 (65) | 1.34 (0.82 - 2.19) | .239 | 1.0 (81) | 1.40 (0.77 - 2.55) | .266 |
| 2020 (Aug) | 1.8 (76) | **2.48 (1.45 - 4.25)** | **.001** | 1.3 (46) | 1.30 (0.78 - 2.15) | .310 | 1.4 (92) | **1.94 (1.10 - 3.41)** | **.022** |
| 2021 (Feb-Mar) | 1.6 (61) | **2.29 (1.31 - 3.98)** | **.003** | 1.3 (49) | 1.37 (0.82 - 2.29) | .233 | 2.0 (105) | **2.86 (1.61 - 5.08)** | **<.001** |
| 2021 (Aug-Sep) | 2.0 (80) | **2.79 (1.65 - 4.73)** | **<.001** | 1.3 (54) | 1.30 (0.79 - 2.14) | .307 | 1.5 (73) | **2.09 (1.14 - 3.83)** | **.017** |
| 2022 (Aug-Sep) | 1.5 (80) | **2.14 (1.26 - 3.64)** | **.005** | 1.0 (49) | 1.03 (0.61 - 1.72) | .921 | 1.4 (77) | **2.08 (1.12 - 3.86)** | **.020** |
| 2023 (Aug-Sep) | 1.5 (66) | **2.08 (1.21 - 3.58)** | **.008** | 1.3 (60) | 1.34 (0.82 - 2.19) | .238 | 1.1 (59) | 1.61 (0.86 - 3.02) | .133 |
| 2024 (Aug-Sep) | 2.7 (115) | **3.79 (2.27 - 6.32)** | **<.001** | 1.5 (64) | 1.51 (0.92 - 2.48) | .106 | 1.6 (99) | **2.36 (1.32 - 4.22)** | **.004** |

*Note: All data except for n are weighted. aOR, 95% CIs, and p values were derived using Stata's post-estimation margins command after fitting separate logistic regression models (one for each outcome), including a survey wave by country interaction term and adjusting for age group, sex, and race/ethnicity. 95% CIs are presented to two decimal places except where they are close to 1, in which case three decimal places are reported. Values in bold indicate p-values <.05.*

**Table 4.** Changes over time in the proportion of youth (16-19 years) who reported past 30-day use of smokeless tobacco, waterpipe/shisha, and heated tobacco within England, Canada, and the US.

|  | England (n=41,327) |  |  | Canada (n=42,499) |  |  | United States (n=45,749) |  |  |
| --- | --- | --- | --- | --- | --- | --- | --- | --- | --- |
|  | %(n) | aOR (95% CI) | P | %(n) | aOR (95% CI) | P | %(n) | aOR (95% CI) | P |
| **Smokeless Tobacco** |  |  |  |  |  |  |  |  |  |
| 2017 (Jul-Aug) | 0.9 (35) | REF | — | 1.1 (40) | REF | — | 2.6 (90) | REF | — |
| 2018 (Aug-Sep) | 1.6 (50) | **3.58 (2.08 - 6.18)** | **<.001** | 1.9 (94) | **1.60 (1.08 - 2.38)** | **.020** | 2.6 (94) | **1.93 (1.26 - 2.93)** | **.002** |
| 2019 (Aug-Sep) | 1.4 (57) | **3.22 (1.91 - 5.44)** | **<.001** | 1.6 (78) | 1.39 (0.91 - 2.11) | .125 | 3.5 (131) | **2.63 (1.72 - 4.01)** | **<.001** |
| 2020 (Feb-Mar) | 2.3 (102) | **5.24 (3.20 - 8.57)** | **<.001** | 1.5 (71) | 1.26 (0.82 - 1.93) | .301 | 2.9 (142) | **2.15 (1.42 - 3.26)** | **<.001** |
| 2020 (Aug) | 1.4 (61) | **3.19 (1.89 - 5.40)** | **<.001** | 1.3 (47) | 1.10 (0.70 - 1.73) | .670 | 1.7 (106) | 1.30 (0.84 - 2.00) | .238 |
| 2021 (Feb-Mar) | 2.0 (84) | **4.64 (2.78 - 7.74)** | **<.001** | 1.3 (53) | 1.12 (0.71 - 1.76) | .631 | 2.5 (125) | **1.84 (1.18 - 2.85)** | **.007** |
| 2021 (Aug-Sep) | 2.9 (111) | **6.55 (4.02 - 10.68)** | **<.001** | 1.8 (74) | **1.57 (1.03 - 2.39)** | **.035** | 2.0 (92) | 1.50 (0.94 - 2.38) | .088 |
| 2022 (Aug-Sep) | 2.1 (99) | **4.83 (2.94 - 7.93)** | **<.001** | 1.3 (56) | 1.14 (0.72 - 1.79) | .577 | 1.4 (80) | 1.03 (0.64 - 1.64) | .915 |
| 2023 (Aug-Sep) | 2.3 (103) | **5.15 (3.13 - 8.47)** | **<.001** | 1.6 (70) | 1.41 (0.93 - 2.16) | .108 | 2.6 (95) | **1.91 (1.19 - 3.08)** | **.008** |
| 2024 (Aug-Sep) | 4.1 (186) | **9.60 (6.01 - 15.33)** | **<.001** | 2.0 (80) | **1.78 (1.16 - 2.71)** | **.008** | 2.5 (142) | **1.84 (1.18 - 2.86)** | **.007** |
| **Waterpipe/Shisha** |  |  |  |  |  |  |  |  |  |
| 2017 (Jul-Aug) | 4.8 (208) | REF | — | 4.1 (170) | REF | — | 4.1 (172) | REF | — |
| 2018 (Aug-Sep) | 4.0 (154) | 0.95 (0.67 - 1.35) | .783 | 3.9 (193) | 0.91 (0.72 - 1.15) | .431 | 3.8 (165) | 1.05 (0.73 - 1.51) | .774 |
| 2019 (Aug-Sep) | 5.8 (216) | **1.42 (1.01 - 1.99)** | **.042** | 4.9 (230) | 1.19 (0.94 - 1.50) | .141 | 4.3 (210) | 1.18 (0.82 - 1.69) | .382 |
| 2020 (Feb-Mar) | 5.7 (270) | **1.40 (1.01 - 1.96)** | **.047** | 4.6 (249) | 1.12 (0.89 - 1.41) | .339 | 3.6 (227) | 0.98 (0.68 - 1.41) | .910 |
| 2020 (Aug) | 4.4 (199) | 1.06 (0.75 - 1.49) | .750 | 3.2 (129) | **0.77 (0.59 - 0.99)** | **.040** | 2.3 (153) | **0.61 (0.41 - 0.88)** | **.009** |
| 2021 (Feb-Mar) | 3.1 (130) | 0.71 (0.49 - 1.03) | .069 | 2.8 (117) | **0.67 (0.51 - 0.88)** | **.004** | 2.8 (155) | 0.73 (0.49 - 1.10) | .133 |
| 2021 (Aug-Sep) | 4.2 (193) | 0.99 (0.71 - 1.40) | .976 | 2.9 (128) | **0.69 (0.53 - 0.89)** | **.005** | 2.0 (132) | **0.54 (0.36 - 0.81)** | **.003** |
| 2022 (Aug-Sep) | 3.8 (173) | 0.91 (0.64 - 1.29) | .605 | 2.8 (131) | **0.65 (0.50 - 0.85)** | **.001** | 2.0 (125) | **0.55 (0.37 - 0.83)** | **.004** |
| 2023 (Aug-Sep) | 4.2 (188) | 0.97 (0.68 - 1.37) | .855 | 2.7 (119) | **0.64 (0.49 - 0.84)** | **.001** | 2.5 (121) | **0.66 (0.44 - 1.000)** | **.050** |
| 2024 (Aug-Sep) | 5.3 (236) | 1.25 (0.89 - 1.76) | .189 | 3.1 (138) | **0.75 (0.57 - 0.98)** | **.033** | 3.1 (170) | 0.83 (0.56 - 1.22) | .338 |
| **Heated Tobacco\*** |  |  |  |  |  |  |  |  |  |
| 2018 (Aug-Sep) | 0.6 (16) | REF | — | 0.6 (27) | REF | — | 1.3 (46) | REF | — |
| 2019 (Aug-Sep) | 0.8 (26) | 1.31 (0.65 - 2.64) | .448 | 1.0 (42) | 1.59 (0.93 - 2.73) | .092 | 1.4 (60) | 1.11 (0.69 - 1.79) | .672 |
| 2020 (Feb-Mar) | 1.4 (64) | **2.38 (1.31 - 4.33)** | **.004** | 0.8 (48) | 1.29 (0.77 - 2.19) | .337 | 1.1 (65) | 0.85 (0.54 - 1.35) | .495 |
| 2020 (Aug) | 1.1 (51) | 1.78 (0.96 - 3.27) | .066 | 0.8 (30) | 1.31 (0.75 - 2.29) | .346 | 1.3 (84) | 1.09 (0.70 - 1.71) | .709 |
| 2021 (Feb-Mar) | 0.9 (36) | 1.55 (0.81 - 2.97) | .184 | 0.9 (37) | 1.46 (0.84 - 2.53) | .177 | 1.1 (66) | 0.84 (0.52 - 1.37) | .490 |
| 2021 (Aug-Sep) | 1.7 (68) | **2.81 (1.55 - 5.12)** | **.001** | 1.0 (44) | 1.63 (0.97 - 2.77) | .067 | 1.0 (59) | 0.78 (0.44 - 1.37) | .391 |
| 2022 (Aug-Sep) | 1.3 (64) | **2.10 (1.15 - 3.85)** | **.016** | 1.0 (45) | 1.54 (0.91 - 2.62) | .110 | 1.0 (53) | 0.78 (0.46 - 1.31) | .343 |
| 2023 (Aug-Sep) | 1.8 (71) | **2.99 (1.64 - 5.46)** | **<.001** | 0.8 (39) | 1.28 (0.75 - 2.19) | .357 | 0.9 (56) | 0.74 (0.42 - 1.30) | .300 |
| 2024 (Aug-Sep) | 2.3 (99) | **3.88 (2.18 - 6.88)** | **<.001** | 1.6 (68) | **2.43 (1.48 - 4.00)** | **<.001** | 0.8 (77) | 0.66 (0.39 - 1.13) | .129 |

*Note: All data except for n are weighted. aOR, 95% CIs, and p values were derived using Stata's post-estimation margins command after fitting separate logistic regression models (one for each outcome), including a survey wave by country interaction term and adjusting for age group, sex, and race/ethnicity. 95% CIs are presented to two decimal places except where they are close to 1, in which case three decimal places are reported. Values in bold indicate p-values <.05. \*Heated tobacco product questions were introduced to the survey in 2018.*

#### Differences in time trends between countries

Supplementary Table S2. shows interactions between country and survey wave. For any product, there was strong evidence of an interaction (F(18,128,301)=8.15, p<0.001): as hypothesised, increases between 2017 and 2024 were greater in England than Canada (aOR=2.73,2.05-3.63) or the US (aOR=2.47,1.93-3.17). There was also strong evidence of interactions for all individual products (p<.05): as hypothesised, increases in England were greater than in Canada and the US for all products except heated tobacco in Canada (all p<.05). Trends were also similar between Canada and the US (all p>.05), except heated tobacco, where increases were greater in Canada (p<.001).

#### Overall country differences

Supplementary Table S3. shows the adjusted country comparisons aggregated across survey waves. Youth in England had higher odds of use than Canada for any product, bidis, waterpipe/shisha, and heated tobacco. Compared with the US, youth in England had higher odds of using bidis, waterpipe/shisha, and heated tobacco but lower odds of using cigarillos, cigars, and smokeless tobacco.

### Has England’s ban on menthol in cigarettes impacted the prevalence of use of non-cigarette tobacco products? (Research Question 3)

Table 5 shows the segmented regression results. Pre-ban, prevalence of use of any non-cigarette tobacco product increased in England (aOR=1.16,1.02-1.31) but declined in Canada (aOR=0.84,0.76-0.93) and the US (aOR=0.82,0.73-0.93), with steeper pre-ban trends in England than Canada and the US (both p<.001). However, use in England did not accelerate after the ban (change in trend: aOR=0.98,0.91-1.06), such that use continued to increase post-ban at a similar rate (post-ban trend: aOR=1.13,1.07-1.20). There was some evidence that post-ban trends differed between England and both Canada and the US (Table 5).

**Table 5.** Segmented regression model estimates of secular (pre-menthol ban) and post-menthol ban change in trends in the proportion of youth (16-19 years) who reported past 30-day use of non-cigarette tobacco products (overall and for each product category), by country, and comparisons between countries.

| Outcome |  | Trends within countries (aOR (95% CI), p-values) |  |  | Comparison of trends between countries (aOR (95% CI), p-values) |  |
| --- | --- | --- | --- | --- | --- | --- |
|  |  | England | Canada | US | England vs Canada | England vs US |
| **Any product** | Pre-ban trend | **1.16 (1.02–1.31), p=.022** | **0.84 (0.76–0.93), p=.001** | **0.82 (0.73–0.93), p=.002** | **1.37 (1.18-1.61), p<.001** | **1.41 (1.23-1.61), p<.001** |
|  | Change in trend | 0.98 (0.91–1.06), p=.632 | **1.07 (1.001–1.15), p=.047** | 1.02 (0.94–1.11), p=.573 | 0.91 (0.82-1.02), p=.094 | 0.96 (0.87-1.06), p=.383 |
|  | Post-ban trend | **1.13 (1.07–1.20), p<.001** | **0.90 (0.86–0.94), p<.001** | **0.84 (0.80–0.89), p<.001** | **1.26 (1.17-1.35), p<.001** | **1.35 (1.27-1.43), p<.001** |
| **Cigarillos** | Pre-ban trend | **1.48 (1.21–1.82), p<.001** | 0.91 (0.78–1.05), p=.198 | **0.81 (0.68–0.96), p=.013** | **1.63 (1.27-2.10), p<.001** | **1.83 (1.48-2.27), p<.001** |
|  | Change in trend | **0.87 (0.77–0.99), p=.037** | 1.03 (0.93–1.14), p=.545 | 1.02 (0.91–1.14), p=.778 | **0.85 (0.72-0.996), p=.045** | **0.86 (0.74-0.99), p=.041** |
|  | Post-ban trend | **1.29 (1.18–1.42), p<.001** | 0.94 (0.88–0.999), p=.046 | **0.82 (0.76–0.89), p<.001** | **1.38 (1.24-1.54), p<.001** | **1.58 (1.43-1.74), p<.001** |
| **Cigars** | Pre-ban trend | **1.53 (1.19–1.95), p=.001** | **0.82 (0.69–0.97), p=.019** | 0.91 (0.73–1.13), p=.386 | **1.87 (1.38-2.52), p<.001** | **1.68 (1.31-2.16), p<.001** |
|  | Change in trend | 0.88 (0.76–1.03), p=.117 | 1.12 (0.99–1.26), p=.065 | 0.97 (0.84–1.13), p=.727 | **0.79 (0.65-0.96), p=.018** | 0.91 (0.76-1.08), p=.279 |
|  | Post-ban trend | **1.35 (1.21–1.50), p<.001** | **0.92-0.85–0.99), p=.018** | **0.88 (0.80–0.98), p=.016** | **1.47 (1.29-1.68), p<.001** | **1.53 (1.36-1.71), p<.001** |
| **Bidis** | Pre-ban trend | **1.49 (1.09–2.04), p=.012** | 1.20 (0.89–1.61), p=.236 | 1.16 (0.84–1.61), p=.355 | 1.25 (0.82-1.91), p=.300 | 1.28 (0.92-1.80), p=.148 |
|  | Change in trend | 0.86 (0.70–1.04), p=.122 | 0.94 (0.77–1.14), p=.524 | 0.92 (0.75–1.13), p=.450 | 0.91 (0.69-1.21), p=.522 | 0.93 (0.73-1.17), p=.523 |
|  | Post-ban trend | **1.27 (1.12–1.47), p<.001** | 1.12( 0.99–1.27), p=.080 | 1.08 (0.93–1.25), p=.329 | 1.14 (0.95-1.37), p=.152 | **1.19 (1.02-1.38), p=.025** |
| **Smokeless Tobacco** | Pre-ban trend | **1.65 (1.24–2.19), p=.001** | 0.97 (0.78–1.20), p=.755 | 0.94 (0.73–1.21), p=.624 | **1.70 (1.19-2.45), p=.004** | **1.75 (1.31-2.34), p<.001** |
|  | Change in trend | 0.90 (0.76–1.07), p=.245 | 1.08 (0.93–1.26), p=.328 | 1.01 (0.85–1.19), p=.948 | 0.83 (0.66-1.06), p=.133 | 0.90 (0.73-1.10), p=.289 |
|  | Post-ban trend | **1.48 (1.30–1.69), p<.001** | 1.04 (0.95–1.15), p=.377 | 0.94 (0.84–1.06), p=.331 | **1.42 (1.21-1.67), p<.001** | **1.57 (1.38-1.79), p<.001** |
| **Waterpipe/ Shisha** | Pre-ban trend | 0.92 (0.78–1.08), p=.304 | 0.88 (0.77–1.01), p=.067 | **0.67 (0.55–0.80), p<.001** | 1.04 (0.84-1.29), p=.704 | **1.37 (1.14-1.67), p=.001** |
|  | Change in trend | 1.04 (0.93–1.16), p=.496 | 0.98 (0.88–1.08), p=.671 | **1.19 (1.04–1.36), p=.012** | 1.06 (0.91-1.24), p=.433 | 0.88 (0.75-1.02), p=.079 |
|  | Post-ban trend | 0.95 (0.89–1.02), p=.191 | **0.86 (0.81–0.91), p<.001** | **0.79 (0.73–0.86), p<.001** | **1.11 (1.01-1.22), p=.033** | **1.20 (1.10-1.31), p<.001** |
| **Heated Tobacco** | Pre-ban trend | **1.95 (1.08–3.53), p=.027** | 1.21 (0.74–1.99), p=.453 | 0.94 (0.59–1.47), p=.772 | 1.61 (0.75-3.49), p=.224 | 2.09 (0.99-4.40), p=.053 |
|  | Change in trend | 0.83 (0.60–1.16), p=.282 | 1.00 (0.74–1.34), p=.975 | 0.94 (0.71–1.26), p=.686 | 0.84 (0.53-1.31), p=.436 | 0.88 (0.57-1.37), p=.582 |
|  | Post-ban trend | **1.63 (1.24–2.14), p<.001** | 1.20 (0.97–1.50), p=.098 | 0.88 (0.72-1.08), p=.221 | 1.35 (0.95-1.92), p=.093 | **1.84 (1.31-2.59), p<.001** |

*Note: All data are weighted. Models are adjusted for age group, sex, and race/ethnicity. 95% CIs are presented to two decimal places except where they are close to 1, in which case three decimal places are reported. Values in bold indicate p-values <.05.*

Similar patterns were observed for cigarillos and cigars. For these products, there was an increasing trend pre-ban in England (cigarillos: aOR=1.48, 1.21–1.82; cigars: aOR=1.53,1.19–1.95) and a declining trend in the US for cigarillos (aOR=0.81, 0.68–0.96) and Canada for cigars (aOR=0.82,0.69–0.97). In England, the trend in cigarillo use slowed after the ban (change in trend: aOR=0.87,0.77–0.99), such that use continued to rise but at a slower rate (post-ban trend: aOR=1.29,1.18–1.42). There was some evidence that the post-ban trend in cigarillo use was higher in England than Canada and the US (Table 5). For cigars, the trend in use did not change after the ban, such that use continued to increase at a similar rate (post-ban trend aOR=1.35,1.21–1.50) and again was greater in England than Canada and the US (Table 5).

For heated tobacco, there was an increasing pre-ban trend in England (aOR=1.95, 95% CI=1.08–3.53) which continued post-ban, with the post-ban trend rising at a greater rate in England than the US (Table 5). For bidis and smokeless tobacco, rising pre-ban trends in England continued post-ban (Table 5). For waterpipe use, there was little evidence for either a pre-ban trend or a change in trends in England and Canada, however, in the US, there was an increase in change in trend (change in trend aOR=1.19,1.04–1.36), though the post-ban trends rose at a greater rate in England than Canada or the US (Table 5).

When assessing use of menthol non-cigarette tobacco products in the years after the ban, as hypothesised, among people who used the products, any menthol (vs. exclusively non-menthol) product and menthol (vs. exclusively non-menthol) cigarillo and cigar use were higher in England compared to Canada (any: 45.3% vs. 38.4%; cigarillos: 40.4% vs. 26.5%; cigars: 28.1% vs. 22.1%), and menthol (vs. exclusively non-menthol) cigarillo use was also higher in England than the US (40.4% vs. 32.4%) (all p<.05; Supplementary Table S4). Among heated tobacco users, menthol (vs. exclusively non-menthol) use was higher in Canada than the US (46.6% vs. 28.2%, p=.006). There was little evidence for any other country differences (Supplementary Table S4).

### Sensitivity analysis

Analyses excluding heated tobacco were similar to the main analyses (Supplementary Tables S4-S7). Trends in menthol cigarette use are provided in Supplementary Table S9)

## DISCUSSION

To our knowledge, this was the first study to assess trends in youth use of non-cigarette tobacco products between 2017 and 2024 in England, Canada, and the US, and whether England’s ban on menthol in cigarettes (but not other tobacco products) impacted prevalence. In England, there was a clear increase in youth use of any non-cigarette tobacco products, particularly cigarillos and cigars, between 2017 and 2024, including, as hypothesised, between late 2020 and 2024. Also as hypothesised, increases in England were greater than those observed in Canada and the US, where use generally remained stable or declined. However, there was little evidence that trends accelerated in England after the menthol ban—while cigarillo and cigar use increased more steeply in England than in Canada (and the US, for cigarillos), the upward trends in England remained similar post-ban, suggesting the ban did not accelerate the extant increase in use.

The hypothesis that any menthol (vs. non-menthol) non-cigarette tobacco product use would be relatively higher in England post-ban was somewhat supported for any products, cigarillos, and cigars, but not for heated tobacco. Specifically, in England, just under half (45%) of youth who used any non-cigarette tobacco product used a menthol product after the menthol cigarette ban, which was higher than Canada (38%) where menthol and other flavours are banned across all tobacco products, but similar to the US where menthol and other flavours are not banned federally (43%). Use of menthol cigarillos was highest in England, and menthol cigar use was higher in England than Canada.

Findings for cigarillo/cigar use suggest some substitution among youth; however, use was already increasing before the menthol cigarette ban, and growth did not accelerate post-ban. This suggests that increasing prevalence may be driven by other factors. For example, cigarillos are exempt from many cigarette regulations in England, meaning that they are cheaper, can be sold in branded packaging, and are available in a range of flavours including menthol and fruit, which may enhance their appeal to youth.^15–20^ More broadly, findings are consistent with overall increases in any nicotine use over the past few years in England,^21,22^ largely attributable to increases in vaping and without an acceleration in the decline of smoking. This may suggest that there is broader uptake of alternative, non-cigarette, nicotine products among youth in England.

Findings have important policy implications. In England, youth use of non-cigarette tobacco products is rising. This is consistent with HM Revenue and Customs figures on cigar use^4^ and earlier data (2020 through 2022) from adult surveys, although more recent data (2022 through 2025) suggest a decline in the proportion of adults using non-cigarette tobacco exclusively (i.e. not alongside cigarettes).^2^ Use of non-cigarette, specifically menthol non-cigarette, tobacco products was also higher in England compared with Canada. In the UK, cigarette policies do not fully apply to non-cigarette tobacco products; for example, menthol and other flavours, standardised packaging, and minimum pack sizes apply to cigarettes but not non-cigarette products. In Canada, such measures apply to a wider range of tobacco products, aligning with lower prevalence of use among Canadian youth. In the US, while menthol and standardised packaging are not federally mandated, the legal age for tobacco sale was raised from 18 to 21 in December 2019,^12^ which aligns with the declines in non-cigarette tobacco product use observed during the study period and is consistent with other data sources.^23^ Therefore, the increases observed in youth use of non-cigarette tobacco products in England but not Canada and the US highlight the need for more comprehensive policies that reduce the appeal and use of these products.

Our data support calls from public health advocates to (a) re-classify cigarillos as factory-made cigarettes for excise and packaging, and (b) extend the menthol and other characterising flavour bans, pack standardisation, and minimum pack size/weight to all tobacco products.^19^ Similar reforms in Canada have been linked to sustained declines in youth combustible tobacco use,^24^ while US local flavour bans show early evidence of reducing flavoured cigar sales.^17^ The proposed UK Smokefree Generation policy will also raise the age of sale for all tobacco products year-on-year, and the 2024-25 Tobacco and Vapes Bill will give the government powers to more comprehensively regulate non-cigarette tobacco products.^25^ These regulations may help England meet its smokefree target.

This study has limitations. First, data collection and implementation of England’s menthol cigarette ban spanned COVID-19, which affected health behaviours, including youth smoking,^26^ and may have confounded trends. Use of waterpipe/shisha, which is more social than use of other non-cigarette tobacco products, showed a clear decline in all three countries around COVID-19. Second, as above, the US federally raised the legal age of sale for all tobacco products to 21 in December 2019^12^ and several US jurisdictions had implemented menthol/flavour bans for tobacco products. Third, samples were not probability based, and survey weights differed between countries: Canada and the US were weighted to reflect national youth smoking trends, while England was not, due to lack of national data among youth aged 16-19. However, smoking and vaping prevalence were similar to national benchmark surveys^14^ and weighting would not have impacted within-country trends. Fourth, assessing past-30-day use means we cannot comment on frequency of use, and assessment of any menthol non-cigarette tobacco product meant that respondents may have used both menthol and non-menthol products although this granularity was not captured in the data.

Fifth, use of some products (e.g., bidis, smokeless tobacco, heated tobacco) was rare, limiting confidence in the findings for these products. Strengths include the use of data from three countries with different policies and a large overall sample that allowed for product-specific analyses.

In conclusion, non-cigarette tobacco product use, including cigarillos, cigars, bidis, smokeless tobacco, and heated tobacco, increased among youth in England between 2017 and 2024. While cigarillo, cigar, and smokeless tobacco use increased more in England than Canada and the US, growth did not appear to accelerate after England’s menthol cigarette ban. Nevertheless, increases in England but not Canada or the US suggest a need for more comprehensive legislation in England that encompasses both cigarettes and non-cigarette tobacco.

## FUNDING

Cancer Research UK provided support for the analyses and contributions to YA, KE, and ET’s salaries (PPRCTAGPJT\100008). Cancer Research UK also supported HTB, SC, and SJ’s salaries (PRCRPG-Nov21\100002). The ITC Youth Survey was supported by a P01 Grant (1P01CA200512) from the US National Institutes of Health (NIH). K.E. is also partially supported by this NIH P01 Grant (1P01CA200512). Waves 3.5, 4.5, and 5 were funded by a contribution from Health Canada’s Substance Use and Addictions Program (SUAP). Oversamples in some US states were supported by the Roswell Park Comprehensive Cancer Center and National Cancer Institute (NCI) (Grant P30CA016056) and by the Roswell Park Alliance Foundation. Oversamples in BC were supported by the British Columbia Ministry of Health / Canadian Cancer Society. The views expressed herein do not necessarily represent the views of any of the funding agencies.

## DECLARATION OF INTERESTS

Authors have never had any financial links to the tobacco or e-cigarette industries or their representatives. JB has received unrestricted research funding from Pfizer and J&J, who manufacture medically licensed smoking cessation medications. D.H. has served as a paid expert witness on behalf of public health authorities in response to legal challenges from tobacco, vaping, and cannabis companies.

## DATA AVAILABILITY

Deidentified study data may be made available on request to researchers who submit a proposal that is approved by the principal investigator. Proposals should be submitted to David Hammond. The analysis code is openly available (https://osf.io/r2y54)..

## Funding

Cancer Research UK provided support for the analyses and contributions to YA, KE, and ET’s salaries (PPRCTAGPJT\100008). Cancer Research UK also supported HTB, SC, and SJ’s salaries (PRCRPG-Nov21\100002). The ITC Youth Survey was supported by a P01 Grant (1P01CA200512) from the US National Institutes of Health (NIH). K.E. is also partially supported by this NIH P01 Grant (1P01CA200512). Waves 3.5, 4.5, and 5 were funded by a contribution from Health Canada’s Substance Use and Addictions Program (SUAP). Oversamples in some US states were supported by the Roswell Park Comprehensive Cancer Center and National Cancer Institute (NCI) (Grant P30CA016056) and by the Roswell Park Alliance Foundation. Oversamples in BC were supported by the British Columbia Ministry of Health / Canadian Cancer Society. The views expressed herein do not necessarily represent the views of any of the funding agencies.

## Declaration of interests

Authors have never had any financial links to the tobacco or e-cigarette industries or their representatives. JB has received unrestricted research funding from Pfizer and J&J, who manufacture medically licensed smoking cessation medications. D.H. has served as a paid expert witness on behalf of public

## Notes

### Author Declarations

Research Ethics Committee of the University of Waterloo gave ethical approval for this work (ORE#21847/31017).

