## Supplementary File for "Trends in youth use of non-cigarette tobacco products in England, Canada, and the US and the impact of England’s menthol cigarette ban on use"

### **SUPPLEMENTARY TABLES**

**Supplementary Table S1. Weighted prevalence (with 95% confidence intervals) of past 30-day use of non-cigarette tobacco products among youth aged 16–19 in England, Canada, and the United States by survey wave (2017–2024).**

|  | **England (n=41,327)** | **Canada (n=42,499)** | **United States (n=45,749)** |
| --- | --- | --- | --- |
|  | **% (95% CI)** | **% (95% CI)** | **% (95% CI)** |
| **Any use of non-cigarette tobacco products** | | | |
| 2017 (Jul-Aug) | 7.4 (6.6–8.2) | 9.2 (8.2–10.3) | 10.2 (9.2–11.3) |
| 2018 (Aug-Sep) | 7.6 (6.7–8.6) | 8.6 (7.8–9.5) | 11.4 (10.3–12.6) |
| 2019 (Aug-Sep) | 8.6 (7.6–9.6) | 9.5 (8.6–10.5) | 11.7 (10.5–13.0) |
| 2020 (Feb-Mar) | 11.0 (10.0–12.0) | 9.0 (8.1–10.0) | 10.2 (9.2–11.2) |
| 2020 (Aug) | 8.4 (7.5–9.3) | 7.4 (6.6–8.3) | 7.6 (6.8–8.6) |
| 2021 (Feb-Mar) | 7.9 (7.0–9.0) | 6.7 (5.9–7.6) | 8.6 (7.6–9.7) |
| 2021 (Aug-Sep) | 9.4 (8.5–10.5) | 7.5 (6.7–8.4) | 7.6 (6.6–8.7) |
| 2022 (Aug-Sep) | 9.8 (8.9–10.8) | 7.3 (6.6–8.2) | 7.9 (6.8–9.2) |
| 2023 (Aug-Sep) | 10.3 (9.3–11.4) | 7.4 (6.6–8.3) | 7.4 (6.4–8.6) |
| 2024 (Aug-Sep) | 11.6 (10.6–12.7) | 7.9 (7.0–8.9) | 7.1 (6.2–8.2) |
| **Cigarillos** | | | |
| 2017 (Jul-Aug) | 2.2 (1.7–2.7) | 4.6 (3.9–5.5) | 6.0 (5.2–6.8) |
| 2018 (Aug-Sep) | 2.5 (2.0–3.2) | 3.7 (3.2–4.3) | 6.8 (5.9–7.8) |
| 2019 (Aug-Sep) | 3.0 (2.5–3.8) | 3.6 (3.0–4.2) | 6.0 (5.2–6.9) |
| 2020 (Feb-Mar) | 3.5 (3.0–4.2) | 4.3 (3.7–5.0) | 5.7 (5.0–6.5) |
| 2020 (Aug) | 3.6 (3.0–4.2) | 4.0 (3.4–4.7) | 4.8 (4.0–5.6) |
| 2021 (Feb-Mar) | 4.0 (3.4–4.9) | 3.6 (3.0–4.2) | 5.1 (4.3–6.0) |
| 2021 (Aug-Sep) | 4.4 (3.7–5.2) | 4.4 (3.8–5.1) | 4.3 (3.5–5.2) |
| 2022 (Aug-Sep) | 4.6 (4.0–5.3) | 3.6 (3.0–4.2) | 4.9 (4.0–6.0) |
| 2023 (Aug-Sep) | 4.4 (3.7–5.1) | 3.9 (3.3–4.6) | 4.6 (3.8–5.6) |
| 2024 (Aug-Sep) | 4.7 (4.0–5.5) | 3.5 (3.0–4.2) | 3.2 (2.7–3.9) |
| **Cigars** | | | |
| 2017 (Jul-Aug) | 1.4 (1.1–1.9) | 2.8 (2.3–3.5) | 3.7 (3.1–4.4) |
| 2018 (Aug-Sep) | 2.0 (1.5–2.5) | 3.2 (2.7–3.7) | 4.3 (3.5–5.1) |
| 2019 (Aug-Sep) | 2.0 (1.5–2.5) | 2.9 (2.5–3.5) | 3.4 (2.8–4.2) |
| 2020 (Feb-Mar) | 2.4 (2.0–3.0) | 2.5 (2.1–3.1) | 3.5 (2.9–4.3) |
| 2020 (Aug) | 2.4 (2.0–3.0) | 2.3 (1.9–2.9) | 2.7 (2.2–3.3) |
| 2021 (Feb-Mar) | 2.5 (2.0–3.2) | 2.1 (1.7–2.6) | 2.9 (2.3–3.6) |
| 2021 (Aug-Sep) | 2.5 (2.0–3.1) | 2.6 (2.1–3.1) | 2.9 (2.3–3.7) |
| 2022 (Aug-Sep) | 2.6 (2.1–3.2) | 2.6 (2.1–3.2) | 2.2 (1.7–2.8) |
| 2023 (Aug-Sep) | 2.9 (2.4–3.6) | 2.6 (2.1–3.2) | 2.9 (2.3–3.8) |
| 2024 (Aug-Sep) | 3.7 (3.1–4.4) | 2.5 (2.0–3.2) | 2.3 (1.8–2.9) |
| **Bidis** | | | |
| 2017 (Jul-Aug) | 1.2 (0.9–1.6) | 1.0 (0.6–1.4) | 1.2 (0.8–1.7) |
| 2018 (Aug-Sep) | 1.2 (0.9–1.7) | 1.1 (0.8–1.4) | 1.7 (1.3–2.2) |
| 2019 (Aug-Sep) | 1.2 (0.9–1.6) | 0.9 (0.7–1.3) | 1.6 (1.2–2.2) |
| 2020 (Feb-Mar) | 2.1 (1.7–2.7) | 1.3 (1.0–1.7) | 1.0 (0.7–1.4) |
| 2020 (Aug) | 1.8 (1.4–2.3) | 1.3 (0.9–1.7) | 1.4 (1.0–1.8) |
| 2021 (Feb-Mar) | 1.6 (1.2–2.2) | 1.3 (1.0–1.8) | 2.0 (1.5–2.6) |
| 2021 (Aug-Sep) | 2.0 (1.6–2.5) | 1.3 (0.9–1.7) | 1.5 (1.1–2.1) |
| 2022 (Aug-Sep) | 1.5 (1.2–1.9) | 1.0 (0.7–1.4) | 1.4 (1.0–2.1) |
| 2023 (Aug-Sep) | 1.5 (1.1–2.0) | 1.3 (1.0–1.7) | 1.1 (0.8–1.6) |
| 2024 (Aug-Sep) | 2.7 (2.2–3.3) | 1.5 (1.1–2.0) | 1.6 (1.2–2.2) |
| **Smokeless Tobacco** | | | |
| 2017 (Jul-Aug) | 0.9 (0.6–1.2) | 1.1 (0.8–1.5) | 2.6 (2.1–3.3) |
| 2018 (Aug-Sep) | 1.6 (1.2–2.1) | 1.9 (1.5–2.3) | 2.6 (2.1–3.2) |
| 2019 (Aug-Sep) | 1.4 (1.1–1.9) | 1.6 (1.3–2.1) | 3.5 (2.8–4.4) |
| 2020 (Feb-Mar) | 2.3 (1.9–2.9) | 1.5 (1.1–1.9) | 2.9 (2.3–3.6) |
| 2020 (Aug) | 1.4 (1.1–1.9) | 1.3 (0.9–1.7) | 1.7 (1.4–2.2) |
| 2021 (Feb-Mar) | 2.0 (1.6–2.6) | 1.3 (1.0–1.7) | 2.5 (1.9–3.2) |
| 2021 (Aug-Sep) | 2.9 (2.3–3.5) | 1.8 (1.4–2.3) | 2.0 (1.5–2.7) |
| 2022 (Aug-Sep) | 2.1 (1.7–2.6) | 1.3 (1.0–1.8) | 1.4 (1.0–1.8) |
| 2023 (Aug-Sep) | 2.3 (1.8–2.8) | 1.6 (1.2–2.1) | 2.6 (1.9–3.5) |
| 2024 (Aug-Sep) | 4.1 (3.5–4.8) | 2.0 (1.6–2.6) | 2.5 (2.0–3.2) |
| **Waterpipe/Shisha** | | | |
| 2017 (Jul-Aug) | 4.8 (4.2–5.5) | 4.1 (3.5–4.8) | 4.1 (3.5–4.9) |
| 2018 (Aug-Sep) | 4.0 (3.3–4.7) | 3.9 (3.4–4.5) | 3.8 (3.2–4.6) |
| 2019 (Aug-Sep) | 5.8 (5.0–6.7) | 4.9 (4.3–5.7) | 4.3 (3.6–5.1) |
| 2020 (Feb-Mar) | 5.7 (5.0–6.5) | 4.6 (4.0–5.3) | 3.6 (3.0–4.3) |
| 2020 (Aug) | 4.4 (3.8–5.1) | 3.2 (2.7–3.8) | 2.3 (1.9–2.8) |
| 2021 (Feb-Mar) | 3.1 (2.5–3.7) | 2.8 (2.3–3.4) | 2.8 (2.2–3.5) |
| 2021 (Aug-Sep) | 4.2 (3.6–5.0) | 2.9 (2.4–3.5) | 2.0 (1.6–2.6) |
| 2022 (Aug-Sep) | 3.8 (3.2–4.5) | 2.8 (2.3–3.3) | 2.0 (1.6–2.6) |
| 2023 (Aug-Sep) | 4.2 (3.5–4.9) | 2.7 (2.2–3.3) | 2.5 (1.9–3.2) |
| 2024 (Aug-Sep) | 5.3 (4.6–6.1) | 3.1 (2.6–3.8) | 3.1 (2.4–3.8) |
| **Heated Tobacco*** | | | |
| 2018 (Aug-Sep) | 0.6 (0.4–1.0) | 0.6 (0.4–0.9) | 1.3 (0.9–1.7) |
| 2019 (Aug-Sep) | 0.8 (0.5–1.3) | 1.0 (0.7–1.5) | 1.4 (1.0–1.9) |
| 2020 (Feb-Mar) | 1.4 (1.1–1.9) | 0.8 (0.6–1.2) | 1.1 (0.8–1.5) |
| 2020 (Aug) | 1.1 (0.8–1.4) | 0.8 (0.6–1.2) | 1.3 (1.0–1.8) |
| 2021 (Feb-Mar) | 0.9 (0.6–1.4) | 0.9 (0.7–1.4) | 1.1 (0.7–1.5) |
| 2021 (Aug-Sep) | 1.7 (1.3–2.2) | 1.0 (0.8–1.4) | 1.0 (0.6–1.5) |
| 2022 (Aug-Sep) | 1.3 (0.9–1.7) | 1.0 (0.7–1.4) | 1.0 (0.7–1.5) |
| 2023 (Aug-Sep) | 1.8 (1.3–2.4) | 0.8 (0.6–1.1) | 0.9 (0.6–1.5) |
| 2024 (Aug-Sep) | 2.3 (1.8–2.9) | 1.6 (1.2–2.0) | 0.8 (0.6–1.3) |
| *Note: All data are weighted. *Heated tobacco product questions were introduced to the survey in 2018.* | | | |

**Supplementary Table S2. Interactions between country and survey wave and comparisons of time trends between countries.**

|  | **Interactions** | **Comparisons of trends between countries** | |
| --- | --- | --- | --- |
|  |  | **England vs Canada aOR (95% CI), p-value** | **England vs US aOR (95% CI), p-value** |
| **Any product** | **F(18,128301)=8.15, p<.001** | **aOR=2.73, 2.05-3.63, p<.001** | **aOR=2.47, 1.93-3.17, p<.001** |
| **Cigarillos** | **F(18,124052)=6.68, p<.001** | **aOR=3.27, 2.07-5.20, p<.001** | **aOR=4.31, 2.96-6.27, p<.001** |
| **Cigars** | **F(18,123981)=5.25, p<.001** | **aOR=5.99, 3.60-9.97, p<.001** | **aOR=4.41, 2.81-6.92, p<.001** |
| **Bidis** | **F(18,123113)=1.92, p=.010** | **aOR=2.51, 1.24-5.07, p=.010** | **aOR=1.60, 0.90-2.87, p=.010** |
| **Smokeless Tobacco** | **F(18,123320)=4.96, p<.001** | **aOR=5.41, 2.90-10.08, p<.001** | **aOR=5.22, 3.12-8.75, p<.001** |
| **Waterpipe/Shisha** | **F(18,123460)=2.05, p=.005** | **aOR=1.67, 1.10-2.55, p=.017** | **aOR=1.52, 1.06-2.16, p=.022** |
| **Heated Tobacco** | **F(16,111417)=2.68, p<.001** | aOR=1.59, 0.74-3.41, p=.230 | **aOR=5.84, 2.68-12.76, p<.001** |

*Note: All data are weighted. Interactions are from logistic regression models adjusted for age group, sex, and race/ethnicity. Values in bold indicate p-values <.05.*

**Supplementary Table S3. Differences between Canada, England, and the United States in the proportion of youth (16–19 years) who reported past-30-day use of non-cigarette tobacco products overall and for each product category between 2017 and 2024.**

|  | **% (n)** | **Canada as reference** | | **US as reference** | |
| --- | --- | --- | --- | --- | --- |
|  |  | **aOR (95% CI)** | **P-value** | **aOR (95% CI)** | **P-value** |
| **Any use (N=128,319)** | | | | | |
| Canada | 8.0 (3,687) | **REF** |  | **-** |  |
| United States | 8.9 (4,772) | **1.14 (1.08-1.21)** | **<.001** | **REF** |  |
| England | 9.2 (4,078) | **1.20 (1.13-1.26)** | **<.001** | 1.05 (0.99-1.11) | .124 |
| **Cigarillos (N=124,070)** | | | | | |
| Canada | 3.9 (1,730) | **REF** |  | **-** |  |
| United States | 5.1 (2,788) | **1.31 (1.21-1.42)** | **<.001** | **REF** |  |
| England | 3.7 (1,566) | 0.95 (0.87-1.03) | .213 | **0.72 (0.67-0.78)** | **<.001** |
| **Cigars (N=123,999)** | | | | | |
| Canada | 2.6 (1,153) | **REF** |  | **-** |  |
| United States | 3.1 (1,622) | **1.15 (1.04-1.27)** | **.006** | **REF** |  |
| England | 2.5 (988) | **0.90 (0.82-1.00)** | **.050** | **0.79 (0.71-0.87)** | **<.001** |
| **Bidis (N=123,131)** | | | | | |
| Canada | 1.2 (529) | **REF** |  | **-** |  |
| United States | 1.4 (767) | **1.23 (1.06-1.42)** | **.005** | **REF** |  |
| England | 1.7 (718) | **1.45 (1.28-1.67)** | **<.001** | **1.19 (1.04-1.36)** | **.011** |
| **Smokeless Tobacco (N=123,338)** | | | | | |
| Canada | 1.5 (663) | **REF** |  | **-** |  |
| United States | 2.4 (1,097) | **1.56 (1.38-1.76)** | **<.001** | **REF** |  |
| England | 2.1 (888) | **1.36 (1.21-1.54)** | **<.001** | **0.87 (0.78-0.98)** | **.020** |
| **Waterpipe/Shisha (N=123,478)** | | | | | |
| Canada | 3.5 (1,604) | **REF** |  | **-** |  |
| United States | 3.0 (1,630) | 0.94 (0.86-1.02) | .143 | **REF** |  |
| England | 4.5 (1,967) | **1.43 (1.32-1.55)** | **<.001** | **1.53 (1.41-1.67)** | **<.001** |
| **Heated Tobacco (N=111,433)** | | | | | |
| Canada | 1.0 (380) | **REF** |  | **-** |  |
| United States | 1.1 (566) | 1.14 (0.96-1.35) | .133 | **REF** |  |
| England | 1.3 (495) | **1.38 (1.18-1.61)** | **<.001** | **1.21 (1.03-1.42)** | **.020** |
| *Note: All data except for n are weighted. Estimates are from logistic regression models adjusting for survey wave, age group, sex, and race/ethnicity. Values in bold indicate p-values <.05.* | | | | | |

**Supplementary Table S4. Differences between Canada, England, and the United States in the proportion of youth (16–19 years) who reported any past-30-day use of menthol non-cigarette tobacco products, among those who reported past 30-day use of each product, overall and for each product category, between August 2020 and 2024.**

|  | **% (n)** | **Canada as reference** | |  | **US as reference** | |
| --- | --- | --- | --- | --- | --- | --- |
|  |  | **aOR (95% CI)** | **p** |  | **aOR (95% CI)** | **p** |
| **Any menthol*** product use (N=5,030)** | | | | | | |
| Canada | 38.4 (511) | **REF** |  |  | **-** |  |
| United States | 42.8 (709) | **1.27 (1.04-1.55)** | **.017** |  | **REF** |  |
| England | 45.3 (864) | **1.42 (1.20-1.67)** | **<.001** |  | 1.12 (0.92-1.35) | .253 |
| **Menthol cigarillos (N=2,518)** | | | | | | |
| Canada | 26.5 (178) | **REF** |  |  | **-** |  |
| United States | 32.4 (294) | 1.27 (0.95-1.71) | .104 |  | **REF** |  |
| England | 40.4 (315) | **1.90 (1.47-2.45)** | **<.001** |  | **1.49 (1.13-1.97)** | **.005** |
| **Menthol cigars (N=1,559)** | | | | | | |
| Canada | 22.1 (97) | **REF** |  |  | **-** |  |
| United States | 23.5 (145) | 1.10 (0.74-1.62) | .637 |  | **REF** |  |
| England | 28.1 (135) | **1.51 (1.07-2.13)** | **.019** |  | 1.37 (0.94-2.01) | .100 |
| **Menthol bidis (N=876)** | | | | | | |
| Canada | 24.5 (57) | **REF** |  |  | **-** |  |
| United States | 31.1 (100) | 1.51 (0.91-2.48) | .109 |  | **REF** |  |
| England | 29.2 (97) | 1.34 (0.86-2.10) | .195 |  | 0.89 (0.57-1.40) | .621 |
| **Menthol smokeless tobacco (N=1,188)** | | | | | | |
| Canada | 32.9 (92) | **REF** |  |  | **-** |  |
| United States | 39.1 (154) | 1.25 (0.82-1.91) | .303 |  | **REF** |  |
| England | 32.1 (155) | 0.99 (0.68-1.42) | .939 |  | 0.79 (0.54-1.15) | .219 |
| **Menthol waterpipe/shisha (N=1,854)** | | | | | | |
| Canada | 26.8 (130) | **REF** |  |  | **-** |  |
| United States | 25.5 (125) | 1.07 (0.74-1.54) | .709 |  | **REF** |  |
| England | 28.4 (227) | 1.17 (0.88-1.56) | .271 |  | 1.09 (0.78-1.54) | .607 |
| **Menthol heated tobacco products (N=743)** | | | | | | |
| Canada | 28.2 (50) | **REF** |  |  | **-** |  |
| United States | 46.6 (103) | **2.25 (1.26-4.03)** | **.006** |  | **REF** |  |
| England | 34.9 (102) | 1.32 (0.80-2.18) | .281 |  | 0.58 (0.34-1.00) | .050 |
| *Note: All data except for n are weighted. Estimates are from logistic regression models adjusting for survey wave, age group, sex, and race/ethnicity. Values in bold indicate p-values <.05. *Menthol-flavoured product questions were introduced in 2021 (wave 5); sample therefore includes waves 5 – 8 only.* | | | | | | |

**Supplementary Table S5. Changes over time in the proportion of youth (16-19 years) who reported past 30-day use of any non-cigarette tobacco products excluding heated tobacco within England, Canada, and the US.**

|  | **England (n=41,327)** | | | **Canada (n=42,499)** | | | **United States (n=45,749)** | | |
| --- | --- | --- | --- | --- | --- | --- | --- | --- | --- |
|  | **%(n)** | **aOR (95% CI)** | ***P*** | **%(n)** | **aOR (95% CI)** | ***P*** | **%(n)** | **aOR (95% CI)** | ***P*** |
| **Any use of non-cigarette tobacco products** | | | | | | | | | |
| 2017 (Jul-Aug) | 7.4 (306) | **REF** | **—** | 9.2 (334) | **REF** | **—** | 10.2 (412) | **REF** | **—** |
| 2018 (Aug-Sep) | 7.5 (281) | **1.41 (1.11-1.81)** | **.006** | 8.4 (423) | 0.87 (0.74-1.03) | .101 | 11.2 (435) | **1.51 (1.20-1.91)** | **<.001** |
| 2019 (Aug-Sep) | 8.5 (322) | **1.62 (1.27-2.06)** | **<.001** | 9.2 (434) | 0.97 (0.82-1.15) | .699 | 11.3 (560) | **1.53 (1.21-1.93)** | **<.001** |
| 2020 (Feb-Mar) | 10.5 (487) | **2.05 (1.62-2.58)** | **<.001** | 8.9 (464) | 0.94 (0.79-1.11) | .449 | 10.0 (619) | **1.33 (1.06-1.67)** | **.015** |
| 2020 (Aug) | 8.1 (380) | **1.54 (1.21-1.95)** | **<.001** | 7.4 (289) | **0.77 (0.64-0.92)** | **.004** | 7.4 (478) | 0.95 (0.75-1.21) | .694 |
| 2021 (Feb-Mar) | 7.8 (315) | **1.45 (1.14-1.85)** | **.003** | 6.4 (279) | **0.66 (0.55-0.8)** | **<.001** | 8.6 (491) | 1.1 (0.86-1.39) | .456 |
| 2021 (Aug-Sep) | 9.3 (421) | **1.77 (1.40-2.24)** | **<.001** | 7.4 (323) | **0.77 (0.64-0.92)** | **.003** | 7.4 (438) | 0.94 (0.73-1.21) | .653 |
| 2022 (Aug-Sep) | 9.5 (459) | **1.85 (1.46-2.33)** | **<.001** | 7.1 (340) | **0.73 (0.61-0.87)** | **<.001** | 7.6 (383) | 0.99 (0.76-1.28) | .918 |
| 2023 (Aug-Sep) | 9.8 (453) | **1.87 (1.48-2.37)** | **<.001** | 7.3 (328) | **0.75 (0.63-0.9)** | **.002** | 7.4 (379) | 0.96 (0.74-1.24) | .739 |
| 2024 (Aug-Sep) | 11.3 (502) | **2.19 (1.74-2.76)** | **<.001** | 7.3 (328) | **0.75 (0.63-0.91)** | **.003** | 7.0 (442) | 0.89 (0.69-1.14) | .351 |
| *Note: All data except for n are weighted. aOR, 95% CIs, and p values were derived using Stata’s post-estimation margins command after fitting separate logistic regression models (one for each outcome), including a survey wave by country interaction term and adjusting for age group, sex, and race/ethnicity. 95% CIs are presented to two decimal places except where they are close to 1, in which case three decimal places are reported. Values in bold indicate p-values <.05.* | | | | | | | | | |

**Supplementary Table S6. Differences between Canada, England, and the United States in the proportion of youth (16–19 years) who reported past-30-day use of any non-cigarette tobacco products excluding heated tobacco between 2017 and 2024.**

|  | **% (n)** | **England as reference** | | **Canada as reference** | |
| --- | --- | --- | --- | --- | --- |
|  |  | **aOR (95% CI)** | **P-value** | **aOR (95% CI)** | **P-value** |
| **Any use (N=126,062)** | | | | | |
| England | 9.0 (3,926) | **REF** |  | **1.19 (1.13-1.26)** | **<.001** |
| Canada | 7.8 (3,542) | **0.84 (0.79-0.89)** | **<.001** | **REF** |  |
| United States | 8.8 (4,637) | 0.97 (0.91-1.02) | .241 | **1.15 (1.09-1.22)** | **<.001** |
| *Note: All data except for n are weighted. Estimates are from logistic regression models adjusting for survey wave, age group, sex, and race/ethnicity. Values in bold indicate p-values <.05.* | | | | | |

**Supplementary Table S7. Segmented regression model estimates of secular (pre-menthol ban) and post-menthol ban change in trends in the proportion of youth (16-19 years) who reported past 30-day use of any non-cigarette tobacco products excluding heated tobacco, by country, and comparisons between countries.**

| **Outcome** |  | **Trends within countries** | | | **Comparison of trends between countries** | |
| --- | --- | --- | --- | --- | --- | --- |
|  |  | **England aOR (95% CI), p-values** | **Canada aOR (95% CI), p-values** | **US aOR (95% CI), p-values** | **England vs Canada aOR (95% CI), p-values** | **England vs US aOR (95% CI), p-values** |
| **Any product** | Pre-ban trend | **1.14 (1.004–1.29), p=.042** | **0.84 (0.76–0.93), p=.001** | **0.82 (0.72–0.92), p=.001** | **1.35 (1.15-1.58), p<.001** | **1.40 (1.22-1.60), p<.001** |
|  | Change in trend | 0.99 (0.91–1.07), p=.742 | 1.06 (0.986–1.14), p=.115 | 1.03 (0.95–1.12), p=.511 | 0.93 (0.84-1.04), p=.196 | 0.96 (0.87-1.06), p=.405 |
|  | Post-ban trend | 1.12 (1.06–1.19), p<.001 | 0.89 (0.86–0.93), p<.001 | 0.84 (0.79–0.89), p<.001 | 1.26 (0.96–1.64) | 1.34 (1.06–1.69) |
| *Note: All data are weighted. Models are adjusted for survey wave, age group, sex, and race/ethnicity. 95% CIs are presented to two decimal places except where they are close to 1, in which case three decimal places are reported. Values in bold indicate p-values <.05.* | | | | | | |

|  | **% (n)** | **England as reference** | |  | | **Canada as reference** | |
| --- | --- | --- | --- | --- | --- | --- | --- |
|  |  | **aOR (95% CI)** | **p** | |  | **aOR (95% CI)** | **p** |
| **Any menthol*** product use (N=4,796)** | | | | | | | |
| England | 43.7 (794) | **REF** |  | |  | **1.36 (1.15-1.61)** | **<.001** |
| Canada | 37.7 (476) | **0.73 (0.62-0.87)** | **<.001** | |  | **REF** |  |
| United States | 41.1 (647) | 0.89 (0.74-1.08) | .253 | |  | 1.22 (0.99-1.49) | .058 |
| *Note: All data except for n are weighted. Estimates are from logistic regression models adjusting for survey wave, age group, sex, and race/ethnicity. Values in bold indicate p-values <.05.* | | | | | | | |

**Supplementary Table S8. Differences between Canada, England, and the United States in the proportion of youth (16–19 years) who reported any past-30-day use of any menthol non-cigarette tobacco products, among those who reported past 30-day use of each product, excluding heated tobacco, between August 2020 and 2024.**

**Supplementary Table S9. Weighted prevalence (with 95% confidence intervals) of any past 30-day menthol cigarette use among youth aged 16–19 who had smoked cigarettes in the past 30 days in England, Canada, and the United States by survey wave (2018–2024 N=15,237).**

| **Country** | **England** | **Canada** | **US** |
| --- | --- | --- | --- |
| 2018 (Aug-Sep) | 42.5 (38.3–46.9) | 30.5 (26.7–34.6) | 55.4 (50.0–60.6) |
| 2019 (Aug-Sep) | 49.9 (45.3–54.4) | 31.1 (26.8–35.7) | 64.3 (58.7–69.5) |
| 2020 (Feb-Mar) | 49.0 (45.3–52.6) | 34.3 (30.2–38.7) | 66.3 (61.1–71.1) |
| 2020 (Aug) | 37.1 (33.1–41.3) | 30.1 (25.3–35.3) | 61.3 (55.4–66.9) |
| 2021 (Feb-Mar) | 36.9 (32.7–41.3) | 25.4 (20.6–31.0) | 61.9 (55.3–68.1) |
| 2021 (Aug-Sep) | 40.1 (35.9–44.3) | 31.9 (27.1–37.0) | 59.8 (52.0–67.1) |
| 2022 (Aug-Sep) | 30.9 (27.8–34.3) | 28.1 (23.5–33.3) | 50.9 (40.3–61.4) |
| 2023 (Aug-Sep) | 35.9 (31.9–40.0) | 39.3 (34.4–44.4) | 60.1 (50.8–68.6) |
| 2024 (Aug-Sep) | 43.8 (40.0–47.6) | 33.7 (29.0–38.7) | 60.0 (54.2–65.5) |
| *Note: All data are weighted. Past 30-day menthol cigarette smoking was not assessed in 2017. The measure used was “In the past 30 days, were any of the cigarettes you smoked flavoured to taste like menthol or mint?”, with response options coded as (1) ‘Yes’ vs. 0 Other (‘No’, ‘Don’t know’, ‘Refused’).* | | | |
